# The influence of common polygenic risk and gene sets on social skills group training response in autism spectrum disorder

**DOI:** 10.1101/2019.12.04.19013888

**Authors:** Danyang Li, Nora Choque-Olsson, Hong Jiao, Nina Norgren, Ulf Jonsson, Sven Bölte, Kristiina Tammimies

## Abstract

Social skills group training (SSGT) is one of the most frequently used behavior interventions in children and adolescents with autism spectrum disorder (ASD). Current evidence suggests that the effects are moderate and heterogeneous. Genetic predisposition could be one of the factors contributing to this heterogeneity. Therefore, we used polygenic risk score (PRS) and gene-set analysis to investigate the association between SSGT response and common variants in autistic individuals. Participants from the largest randomized clinical trial of SSGT in ASD to date were selected for genotyping. Polygenic risk scores (PRSs) for ASD, attention deficit hyperactivity disorder (ADHD), and educational attainment (EA) were calculated, and their associations with the intervention outcome at post-intervention and follow-up were tested using mixed linear model. In addition, thirty-two gene sets within five categories (synaptic, glial, FMRP, glutamate, and mitochondrial) were selected to evaluate their role in the intervention outcome. Individuals with higher PRSs for ASD and ADHD had inferior response after SSGT. After multiple test correction, significant results were kept for higher ADHD PRS at follow-up (β = 6.67, p = 0.016). Five gene sets within synaptic category showed modest association with reduced response to SSGT in ASD. Taken together, we provided preliminary evidence that genetic liability calculated using PRS and common variants in synapse gene sets could influence the outcome of SSGT. Our results hold promise for future research into the genetic contribution to individual response to ASD interventions, and should be validated in larger cohorts.

## Introduction

The response to behavioral interventions varies between individuals with the same neurodevelopmental and psychiatric disorders. One of the factors contributing to this heterogeneity might be genetic predisposition. Genome-wide association studies (GWAS) have been used to pinpoint common variants, with mostly small effects, associated with disorders or personality traits across a genome. For behavioral interventions, the size of available cohorts with detailed outcomes is limited, and to date no significant variants have been identified to be associated with outcome for any specific interventions using GWAS. However, the use of polygenic risk score (PRS), an aggregate measure of the cumulative effects of single nucleotide polymorphisms (SNPs) derived from GWAS, has provided promising results in psychiatry both for behavioral and pharmacological treatments^1–5^. For instance, PRS has been studied to predict the response to cognitive behavior therapy in major depressive disorder^2^. In addition to PRS, gene-set analysis can be utilized to group multiple genetic variants in genes and further to related gene sets to unravel biological process and cellular functions related to intervention responses. Relevant gene set associations have been identified for interventions in psychiatry^6–10^. For example, genetic variations in genes underlying glutamatergic or NMDA neurotransmission have been implicated in short-term antipsychotic medication efficacy in schizophrenia and calcium signaling pathway has been indicated in response to selective serotonin reuptake inhibitors in obsessive-compulsive disorder^8,10^.

Autism spectrum disorder (ASD) is a neurodevelopmental disorder characterized by impairments in social communication and interaction, together with restricted, repetitive behaviors^11^. ASD commonly co-exists with other neurodevelopmental and psychiatric disorders such as attention-deficit hyperactivity disorder (ADHD), anxiety, depression, and intellectual disability^12^. The genetic knowledge in ASD has increased rapidly in recent years. For numbers of the individuals with ASD, rare genetic variants, such as loss-of-function variants and copy number variations (CNVs) in specific genes and loci, can indicate a molecular etiology^13^. In addition, common variants have been shown to be of causal significance in the disorder^14–16^. Although there are only few genome-wide significant SNPs identified for ASD, the cumulative polygenic variation summarized by PRS has shown to be predictive of ASD and autistic traits in the general population^17,18^. Studies have also indicated shared genetic liability of polygenic risk in ASD and psychiatric disorders^19,20^. Educational attainment (EA), defined as the highest degree of education, also has a confirmed genetic correlation with ASD^21^ and academic achievement has been linked to social skills^22^. Both rare and common genetic variations in ASD have shown to converge to specific gene-sets such as synaptic formation and targets of the Fragile-X mental retardation protein (FMRP)^23,24^. To date, there are no studies investigating how the already implicated common genetic variation in ASD would relate to intervention outcomes in autistic individuals.

Social skills group training (SSGT) is one of the most frequently used behavioral interventions in ASD, aiming to alleviate social communication difficulties in verbal individuals within normative intellectual range in a group setting. The largest randomized controlled trial (RCT) of SSGT (“KONTAKT”) to date, conducted by our center in Sweden, included children and adolescents with ASD and at least one common neurodevelopmental or psychiatric comorbidity^25^, such as ADHD, anxiety and depression. SSGT as an add-on to standard care was found to have a small to moderate effect compared to standard care only, with significant effects on the primary outcome limited to adolescents (13-17 years) and females^25^.

We recently showed that autistic individuals who were carriers of clinically significant and rare genic CNVs larger than 500kb had significantly inferior outcomes after SSGT within the RCT^26^. Here, we expand our genetic investigations to test the association between SSGT intervention response in ASD and common variants using PRS and gene-set analysis. We hypothesized that PRSs for ASD, ADHD, and EA, as well as common variants within known ASD gene sets, would influence the response to SSGT. Additionally, we tested if there was a significant correlation with the clinical measures of SRS, ADHD diagnosis and IQ in our clinical ASD cohort with the selected PRS. To the best of our knowledge, this is the first study to evaluate the influence of common variants on SSGT response in autism or any other neurodevelopmental disorders. Our results will provide further evidence of potential use of genetic profiles to predict individual-level outcomes for ASD interventions.

## Methods

### Study individuals

The original multicenter, randomized pragmatic RCT of SSGT (“KONTAKT”) recruited participants from 13 child and adolescent psychiatry outpatient units in Sweden between August 2012 and October 2015. A detailed description of the inclusion and exclusion criteria have been earlier described by Choque-Olsson et al^25^. In short, 296 children (7–12 years) and adolescents (13–17 years) with a diagnosis of autism, atypical autism, Asperger syndrome, or pervasive developmental disorder not otherwise specified using ICD-10 criteria were included in the trial^27^. All participants had full-scale IQ > 70 according to the Wechsler Intelligence Scale for Children and at least one common comorbid psychiatric diagnosis of ADHD, depression, or anxiety disorder according to ICD-10^27,28^. During the 12-weeks trial, the standard care group (n=146) received any ongoing support or intervention, and the remaining 150 participants were included in the active SSGT condition. The parent-reported Social Responsiveness Scale (SRS) assessing autistic-like traits was used as the primary intervention outcome measure. The SRS is a 65-item Likert-type scale generating totals scores ranging between 0 and 195, with higher score indicating greater autism trait severity^29^. SRS was recorded for each individual at baseline (pre-intervention), 12 weeks (post-intervention), and 3-month after the end of the intervention (follow-up).

Participants who contributed saliva samples and had the primary outcome measure recorded at either post-intervention or follow-up were included in this study. After selection, clinical data and samples from 207 participants (SSGT group: 105, standard care group: 102) were used for genotyping as described in elsewhere^26^.

### Ethics

Written informed consent from the parents or legal guardians and verbal consent from the children and adolescents was collected. All the protocols and methods in this study were in accordance with the Declaration of Helsinki. The trial and sample collection were approved by the ethical review board in Stockholm (Dnr 2012/385-31/4) and the clinical authorities of the two involved counties. The trial was registered online (NCT01854346).

### Genotyping and quality control

DNA collection, extraction, and genotyping procedures are described elsewhere in detail^26^. In short, the genotyping was done on the Affymetrix CytoScan™ HD microarray platform, containing 743 304 SNP probes, in two separate batches. Data from genotyping were transformed from Affymetrix .CEL format to .tped format using “Affy2sv” package v1.0.14 in R. Position of each SNP was located based on microarray annotation reference file (version NA32.3). Quality control (QC) of the data using PLINK v1.90 was performed on per-individual basis within each genotyping batch to remove poorly genotyped individuals and on per-marker QC to exclude low-quality markers following a published protocol^30,31^. Individuals with discordant sex, heterozygosity rate > 3SD, individual genotype failure rate > 0.03, and relatedness were removed. Ancestry of the participants was estimated using principle component analysis (PCA) in EIGENSOFT v7.2.1. We restricted our analyses to European ancestry.

Thus, individuals were excluded based on two largest principle components (principle component 1 < 0.00, principle component 2 < 0.05). Additionally, the largest four PCAs were added in the statistical model to adjust for ancestry. As no batch effects were detected, the qualified data were combined to clean low-quality markers with following criteria: minor allele frequency < 0.05, Hardy-Weinberg equilibrium < 1e-06, individual missingness < 0.1, and marker missingness < 0.05.

### Imputation

We used the 1000 Genomes phase III haplotype data including all ancestries as a reference genome. SNPs passing QC were separated into autosomes, and haplotypes were inferred based on reference panel using SHAPEIT v2^32^. For each phased autosome, imputation was performed in 5Mb windows using IMPUTE2 v2.3.2^33,34^. All imputed regions were combined for post-imputation QC. Imputed SNPs were filtered using following metrics: info score < 0.8, minor allele frequency < 0.05, Hardy-Weinberg equilibrium < 1e-06, marker missing rate < 0.05, and individual missing rate < 0.1. SNPs were intersected together after post-imputation QC using both SNPTEST v2.5.5 and PLINK v1.90^31,35^.

### Polygenic Risk Score calculation

To calculate the PRSs for ASD, ADHD, and EA, we used the latest GWAS summary results from Psychiatric Genomics Consortium^16,36,37^. To minimize different population effects, we included only individuals with European ancestry from the GWAS reference samples. The estimated odds ratio and P-value of each SNP allele were used from each reference set. SNPs in both reference and our in-house data were pruned using clumping with a cutoff of r^2^ >= 0.1 within 500 kb window. PRS was calculated based on independent SNPs using five P-value thresholds (Pt) (< 0.01, < 0.05, < 0.1, < 0.5, < 1) selected from three reference sets, with higher Pt incorporating more SNP effects. Allele numbers at each SNP (0, 1, 2), weighted by the natural logarithm of the allelic odds ratio were summed to calculate an accumulative effect across the genome^1,38^. PRS was then standardized (mean = 0, SD = 1) to test for association. All calculations were conducted using PRSice v2.1.4^39^.

### Gene sets generation

Reference gene sets were obtained based on a previously published study^24^. Thirty-two gene sets within five categories: synaptic (1047 genes), glial (240 genes), FMRP (1809 genes), glutamate (156 genes), and mitochondrial (132 genes) were included. SNPs were annotated to genes based on European population from 1000 genomes and gene locations, Build 37.

### Statistical analysis

#### PRS association

We used mixed linear model (MLM) to identify the effect of PRS on the SSGT response. Three time-points (pre-, post-intervention, and follow-up), as well as two study groups (SSGT and standard care) were included to test the interaction effect of PRS*time*intervention. Based on our previous studies^25,26^, factors associated with inferior intervention outcomes were younger age (children), male gender, and CNV carrier status (large size CNVs (> 500kb) and clinically significant CNVs). Therefore, age group, gender, and population stratification of the four largest PCAs were added as fixed factors in the model. Additionally, the clinical centers and each individual were used as random factors. Carrier status of large size CNVs and clinically significant CNVs were also added as fixed factors in separate models comparing with the model without CNV adjustment. Marginal R^2^ for fixed effect and conditional R^2^ for fixed and random effects were assessed to explain the role of PRS on the variance of treatment outcome. For each phenotype on each Pt, both marginal and conditional R^2^ were derived from the difference between model with PRS and model without PRS, performed by “MuMIn” package v1.43.6 in R^40^. Beta and 95% confidential interval (CI) were estimated from MLM to evaluate the interaction effect of PRS with SSGT and standard care at different times. After false-discovery rate multiple corrections among three PRSs, original P-values under 0.0167 were considered significant^41^. In addition, two-sided student’s t-test and Pearson correlation were used to examine the correlation between PRS and closely related characteristics, including PRS for ADHD and ADHD comorbidity status, PRS for EA and IQ level, and PRS for ASD and ASD severity (SRS score at pre-treatment). All analyses were performed in R v3.4.2.

#### Gene sets association

MAGMA v1.0.6 was used to perform gene and gene-set analysis^42^. Linear regression was chosen to identify the effects of common variation within specific gene sets on intervention outcome. The changes of SRS score between post-intervention or follow-up and pre-intervention were used as regression outcomes. Age, gender, four largest PCAs and CNV carrier status (large size CNVs and clinically significant CNVs) were added in the model as cofactors. The estimated effect size of competitive test and P-value on each gene set were obtained followed by multiple testing correction with positive effect size indicating a decreased effect of the SSGT response.

## Results

### Sample characteristics

Samples of 188 participants passed genotyping QC, of which 99 belonged to active SSGT. When comparing the individuals of this RCT subgroup with the total cohort from the RCT^25^, we did not detect any differences in characteristics between these individuals and those not passing genotyping QC (Supplementary Table 1). The final genotype data sets consist of 539 106 SNPs after genotyping marker QC and 5 126 694 SNPs after imputation QC.

### Correlation between PRS and related characteristics

The correlation between ADHD comorbidity status and PRS for ADHD, pre-intervention SRS and PRS for ASD, as well as IQ and PRS for EA were tested (Supplementary Figure 1). No correlation was observed in ASD PRS and baseline SRS (Supplementary Figure 1A). However, autistic participants with ADHD comorbidity had higher ADHD PRS compared with participants without ADHD (Pt = 0.1, p = 0.010; Pt = 0.5, p = 0.0075; Pt = 1.0, p = 0.0077) (Supplementary Figure 1B), and IQ was positively correlated with PRS for EA (Pt = 0.05, p = 0.038; Pt = 0.1, p = 0.024; Pt = 0.5, p = 0.014; Pt = 1.0, p = 0.012) (Supplementary Figure 1C).

### Proportion of variance in SSGT outcome explained by PRS

For the model examining the effect of PRS for ASD, the largest explained marginal and conditional variance in the SSGT outcome was Pt 0.5 when controlling for the clinically significant CNVs (Figure 1). We detected only small differences in the explained variance when testing for ADHD PRS at all Pt (Figure 1). For EA PRS, increasing values of marginal and conditional R^2^ occurred with higher Pt, in which Pt 1.0 explained the most variance. All three tested models showed similar results (Figure 1, Supplementary Figure 2).

**Figure 1.**
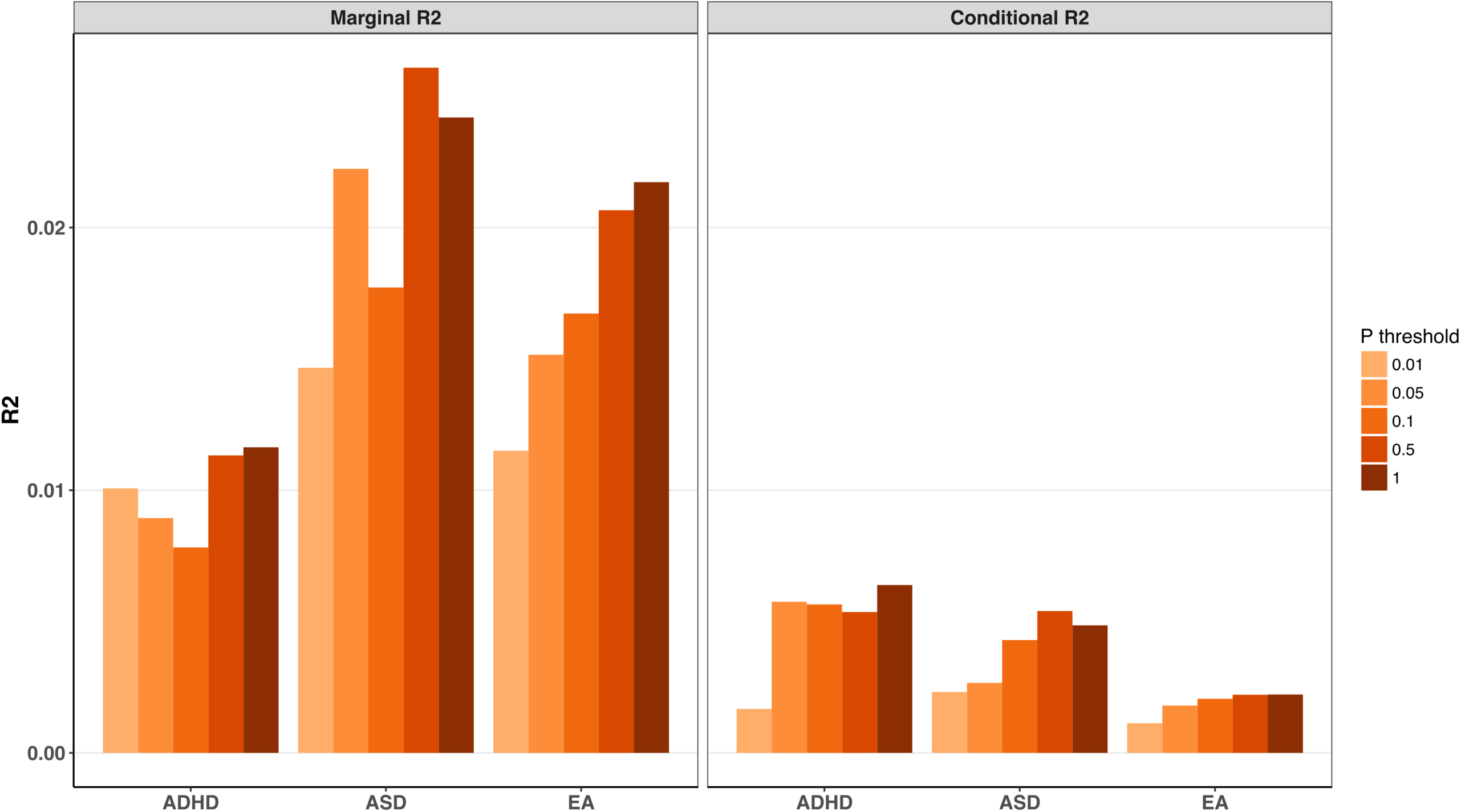
Proportion of variance explained (R^2^) by polygenic risk scores (PRSs) of autism spectrum disorder (ASD), attention deficit hyperactivity disorder (ADHD), and education attainment (EA) in social skill group training (SSGT) derived using five P-value thresholds (< 0.01, < 0.05, < 0.1, < 0.5, < 1) in the model adjusted for clinical significant rare copy number variation (CNV). Marginal R^2^ and conditional R^2^ were calculated representing the variance explained by only fixed effects as well as the sum of fixed and random effects.

### Association between PRS and SSGT outcome

The PRS calculated from the Pt which had the highest explained variance for each phenotype is shown as main result (Figure 2). Associations for all Pts in three models are shown in Supplementary Table 2. Inferior outcomes after SSGT were implicated in individuals with higher PRS for ASD on Pt 0.5 (β = 6.47, p = 0.019) and Pt 1.0 (β = 5.93, p = 0.031) at follow-up. However, the associations were not significant after multiple testing correction. For ADHD PRS, participants with higher score also improved less from SSGT on Pt 0.05 (post-intervention: β = 6.22, p = 0.019, follow-up: β = 6.93, p = 0.012), Pt 0.1 (post-intervention: β = 6.22, p = 0.017, follow-up: β = 5.75, p = 0.036) at both post-intervention and follow-up and Pt 0.5 (β = 6.00, p = 0.030), Pt 1.0 (β = 6.67, p = 0.016) at follow-up. After multiple testing correction, significant association remained in the follow-up for ADHD PRS score at Pt 0.05 and Pt 1.0. No significant effects were found for EA PRS.

**Figure 2.**
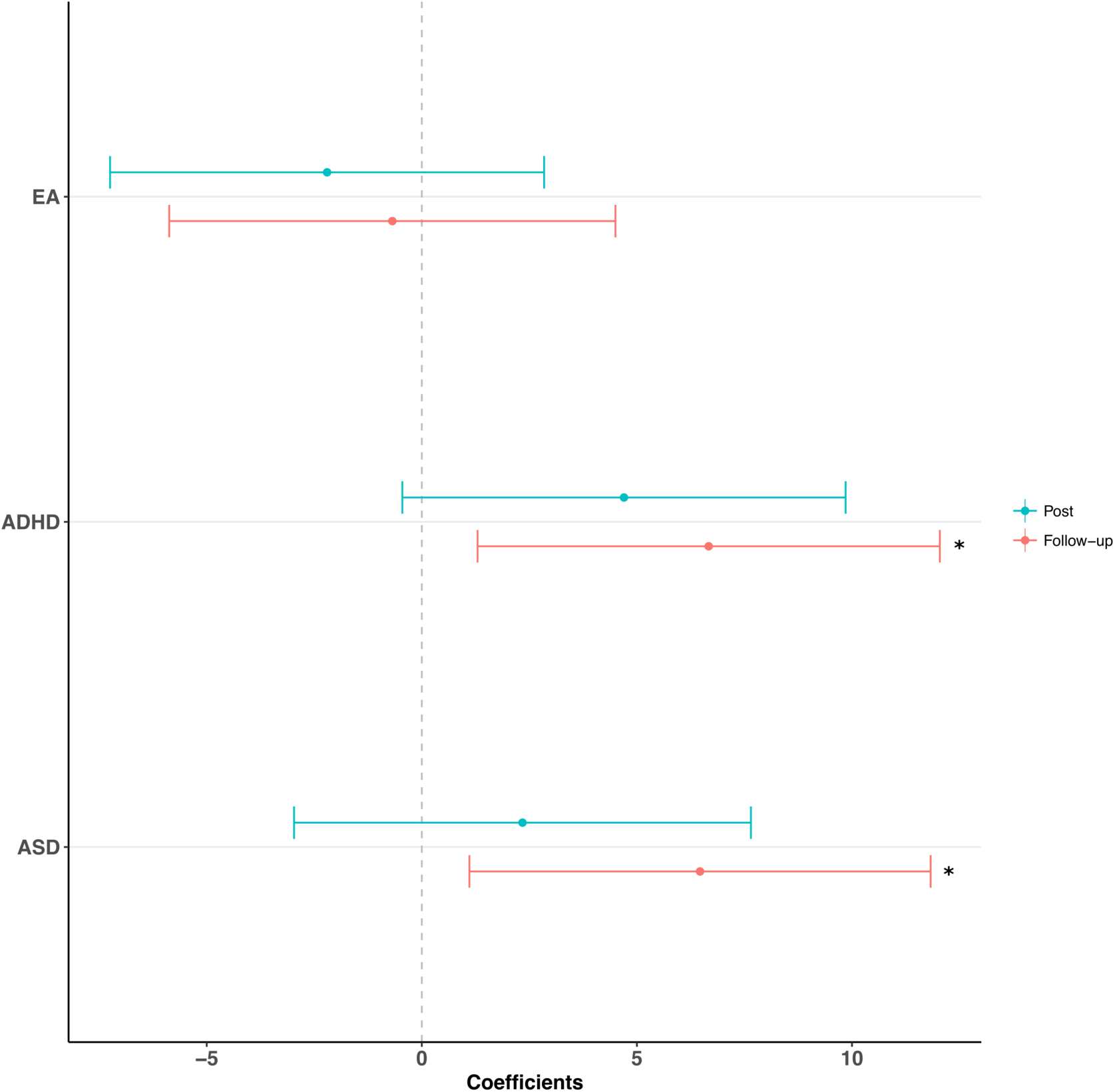
Association of polygenic risk score (PRS) for autism spectrum disorder (ASD), attention deficit hyperactivity disorder (ADHD), and educational attainment (EA) at post-intervention and follow-up in social skill group training (SSGT). Correlation coefficients are shown with 95% confidence intervals. Different P-value thresholds which have highest explained variance for each PRS were included in the model (ASD: Pt = 0.5, ADHD: Pt = 1, EA: Pt = 1). Large size rare copy number variations (CNVs) (>500kb) and clinically significant rare CNVs were added as cofactors respectively in the model. *: P < 0.05.

### Gene sets association with SSGT response

We tested if common genetic variation in five gene set groups and 31 ASD related gene sets could explain SSGT outcome using competitive gene-set analysis. For the outcome at post-intervention, four gene sets showed modest effect to cause significant inferior outcome: intracellular signal transduction (large size CNVs: β = 0.204, p = 0.0027; clinically significant CNVs: β = 0.202, p = 0.0029), cell adhesion and trans-synaptic signaling (large size CNVs: β = 0.255, p = 0.0071; clinically significant CNVs: β = 0.247, p = 0.0090), excitability (large size CNVs: β = 0.268, p = 0.017; clinically significant CNVs: β = 0.247, p = 0.026), GPCR signaling (large size CNVs: β = 0.255, p = 0.023; clinically significant CNVs: β = 0.252, p = 0.024), all belonging to synaptic group (Table 1). In follow-up, only RNA and protein synthesis, folding and breakdown (RPSFB) (large size CNVs: β = 0.184, p = 0.030; clinically significant CNVs: β = 0.185, p = 0.030) in synaptic group showed similar effect (Table 1). However, none of the gene set results remained significant after multiple testing correction. The results for other gene sets did not reach nominal significance (Supplementary Table 3).

**Table 1.** Effect of most significant gene sets (P < 0.05) on social skill group training (SSGT) at post-intervention and follow-up stratified by clinically significant rare copy number variations (CNVs) and large size (> 500kb) rare CNVs

| Gene sets | Gene set groups | Number of genes | Clinically significant CNVs |  |  |  | Large size CNVs |  |  |  |
| --- | --- | --- | --- | --- | --- | --- | --- | --- | --- | --- |
|  |  |  | Beta | Se | P | Corrected P | Beta | Se | P | Corrected P |
| Post-intervention |  |  |  |  |  |  |  |  |  |  |
| Cell adhesion and trans-synaptic signaling | Synaptic | 74 | 0.247 | 0.104 | 0.0090 | 0.2340 | 0.255 | 0.104 | 0.0071 | 0.1988 |
| Excitability | Synaptic | 55 | 0.247 | 0.127 | 0.0262 | 0.5383 | 0.268 | 0.127 | 0.0174 | 0.4110 |
| GPCR signaling | Synaptic | 40 | 0.252 | 0.128 | 0.0244 | 0.5142 | 0.255 | 0.128 | 0.0231 | 0.5011 |
| Intracellular signal transduction | Synaptic | 138 | 0.202 | 0.073 | 0.0029 | 0.0837 | 0.204 | 0.073 | 0.0027 | 0.0832 |
| Follow-up |  |  |  |  |  |  |  |  |  |  |
| RPSFB | Synaptic | 62 | 0.185 | 0.098 | 0.0303 | 0.5871 | 0.184 | 0.098 | 0.0305 | 0.5928 |

Abbreviations:
GPCR: G-protein-coupled receptor; RPSFB: RNA and protein synthesis, folding and breakdown

## Discussion

In this study, by calculating PRS and analyzing related gene sets, we tested if different subsets of common variants and biological gene groups would influence the outcome of SSGT. We showed that a higher common genetic variant load for ADHD and ASD was negatively associated with the SSGT response in autistic individuals, albeit with modest effects compared to rare CNVs impact we have shown before^26^. However, the effect of PRS remained even when we controlled for rare CNV carrier status of the individuals suggesting an independent role of PRS on the intervention outcome. We also demonstrated significant correlations between PRS for ADHD and ADHD comorbidity and PRS for EA and IQ level in our study cohort. Additionally, we showed suggestive evidence that genes involved in synaptic functions could be important for modulating the response to SSGT, as several gene sets important for synaptic functions showed nominal significance to influence the SSGT outcome.

Given the high heterogeneity of responses to SSGT among individuals with ASD, it is important to find factors influencing the outcome, which could be used to tailor the interventions for each individual. Our results provided intriguing evidence that in addition to the rare high impact CNVs, the polygenic contribution of common variants could have a moderating role on SSGT outcome, especially the ADHD PRS. Highly shared genetic heritability between ASD and ADHD has been reported in family studies^43,44^, and genetic overlap between the two neurodevelopmental conditions is evident from genome-wide correlation analysis^21,45,46^. A recent study has identified that PRS for ADHD can be used to predict ASD-related measures such as pragmatic language abilities^47^. Compared to ASD alone, having ASD and ADHD symptoms were confirmed to be associated with greater impairments in socialization adaptive skills in clinical presentation^48^. Based on genetic sharing between the two conditions, our results highlight the connection between ADHD genetic information and ASD intervention effects. Furthermore, the correlation between PRS and ADHD comorbidity indicates that ADHD PRS could be used to evaluate liability to ADHD in individuals first diagnosed with ASD.

Compared to the results of PRS for ADHD, we found only indicative association between PRS for ASD and intervention outcomes. Currently, it is not known if the same genetic factors would influence the risk of developing ASD and intervention response measured here. In other disorders such as major depressive disorder (MDD), studies have found variants associated with MDD can be either positive or negative for the outcome of antidepressants^49^, and the results of PRS as a predictor of treatment outcome are inconsistent^50,51^. Further studies should clarify how ASD PRS might modulate intervention outcomes.

We did not find any association between PRS for EA and SSGT intervention response. Studies using genetic correlation analysis corroborated interrelated results between ASD and years of education and college attainment^21,52^. Some PRS studies suggest that PRS for ASD is associated with EA^52^, and EA PRS is associated with lower externalizing behaviors^53^, but no study has shown the prediction of autistic traits or ASD-related interventions using PRS for EA. As expected, we found a positive correlation between PRS EA and IQ in our sample, which is in accordance with other findings in the general population^54^.

Interestingly, the inferior outcome for individuals with higher PRS for ASD and ADHD seemed to be particularly pronounced at follow-up. A similar tendency was previously also seen in individuals with clinically significant rare CNVs^26^. This suggest that additional care during or after SSGT treatment should be considered for individuals with higher genetic risk, as to alleviate the genetic influence on the intervention outcome. Recently, a long version of SSGT intervention (24-week) was conducted resulting in larger positive effects compared to the 12-week intervention^55^. Further studies should investigate if genetic effects remain when the intervention dose is increased.

Although no robust association after multiple testing corrections were shown for the gene-set analysis, we found suggestive evidence that genes necessary for synapse formation and maintenance may influence SSGT response. Since synapse formation and synaptic plasticity have been indicated as one of the key neuronal mechanisms in ASD, differences in these key steps in brain development could possibly play a major role in how individuals respond to interventions^56^. Some aspects of treatment-induced behavior improvement are also related to brain plasticity changes in psychiatric disorders. For instance, significant increases of grey matter in left hippocampus and left amygdala correlated with the degree of improved cognition are found in early-onset schizophrenia individuals after two years of social skills group therapy and cognitive remediation^57^.

We acknowledge the limitations of our study. First, the sample size from RCT is limited, which restricted the precision of our analyses and precluded in-depth analysis of complex pathways and single variants. Secondly, the analyzed sample included individuals with normative IQ and common neurodevelopmental and psychiatric comorbidities, limiting the generalizability across the total population of autistic individuals. Finally, studies have shown loci harboring common alleles are also enriched for rare variants with large effects from whole-exome sequencing in psychiatric disorders^58^. Although we have adjusted for the carrier status of rare CNVs, other rare variants were not controlled for. In the future, more genetic information can be combined to better understand their effect on SSGT outcome.

### Conclusion

This is the first study showing that common polygenic risk, especially for ADHD, is associated with SSGT response in ASD individuals, and gene sets from synaptic groups may play a potential biological role in SSGT response. Replications including larger sample size and combination of more genetic and clinical factors are needed to further clarify the genetic influences and mechanisms behind individual intervention responses.

### Web resources

1000 Genomes Phase III haplotypes reference data: https://mathgen.stats.ox.ac.uk/impute/impute_v2.html#reference Psychiatric Genomics Consortium: https://www.med.unc.edu/pgc/ Gene locations from the NCBI site and SNP locations from 1000 genomes Phase 3 European population Build 37 for MAGMA: https://ctg.cncr.nl/software/magma

## Data Availability

Due to the legal restriction in the Swedish law we are not allow to share the raw genetic data used in the study

## Acknowledgments

We thank the children, adolescents, and parents who participated in the study. Christina Coco MSc, Oskar Flygare, MSc, Anders Görling MSc, and Kerstin Andersson MSc are acknowledged for their work in collecting the data and samples during the RCT. We also thank Ielyzaveta Rabkina and Sofia Stamouli for the help with handling the DNA samples and genetic data. The authors are also thankful to the leads of child and adolescent psychiatry (Olav Bengtsson, MD, Paula Liljeberg, MD, Charlotta Wiberg Spangenberg, MSc, Peter Ericson, MSc, Karin Forler, MSc, Alkisti Nikolayidis Linderholm, MSc, all of Stockholm County Council), PRIMA Järva child and adolescent psychiatry (MaiBritt Giacobini, MD, PhD), and child and adolescent habilitation services (Lars Kjellin, PhD, Moa Pellrud, MSc, of Örebro County Council) for organizational support. We would like to acknowledge Martin Becker and Ielyzaveta Rabkina for their contributions to the genetic work. The DNA extraction was done at the KI Biobank. The genotyping was done at Aros Applied Biotechnology in Denmark. The computation resources were provided by SNIC through Uppsala Multidisciplinary Center for Advanced Computational Science (UPPMAX). We also want to thank the Swedish Bioinformatic Advisory program for providing bioinformatic consultation for data analysis.

## Contributions

D.L and K.T conceived and planned the presented study. D.L performed the analysis with supervision from N.N, and K.T. H.J. provided overall suggestion and discussion for the analyses. S.B and N.C-O conceived the original clinical trial. S.B is responsible for the clinical data and provided expertise of the clinical measures together with N.C-O, and U.J. D.L and K.T wrote the paper with input from all authors. All authors approved the final version of the paper.

